# Epigenetic Age Prediction Remains Stable Across Common Variants and Diverse Ancestries

**DOI:** 10.1101/2025.10.26.25338818

**Authors:** Turki M. Sobahy, Estefania Huergo, Jesper Tegner, Vincenzo Lagani, David Gomez-Cabrero

## Abstract

Epigenetic clocks, based on DNA methylation profiles at CpG sites, are widely recognized as reliable biomarkers of biological aging. However, common single-nucleotide polymorphisms (cSNPs), genomic variants that can overlap CpG sites, may affect DNA methylation profiles in ways that potentially interfere with the accuracy of epigenetic clocks. Moreover, because the prevalence of cSNPs varies across populations, such cSNP-CpG overlaps may differentially affect the age predictions of epigenetic clocks in diverse cohorts. Here, we present the first systematic cross-ancestry evaluation of cSNP robustness in the epigenetic clock, examining how cSNP-CpG overlaps affect the performance of epigenetic clocks across nine major genomic ancestry groups. We employed three complementary strategies: (a) testing whether cSNP-CpG overlaps are overrepresented in established epigenetic clocks or particular populations, (b) evaluating whether overlapping CpG sites correspond to the most influential aging predictors within clock models, and (c) simulating the effects of cSNP-associated methylation changes on predicted biological age. Our findings indicate that cSNP-CpG overlaps are not enriched among the CpG sites used in current epigenetic clocks, nor do they tend to involve the most influential sites. Furthermore, our simulation analysis revealed that current epigenetic clocks appear robust to cSNP-related methylation variations. Our findings underscore the overall stability of current epigenetic clocks, even in the presence of population-specific cSNP-CpG overlaps that are known to affect DNA methylation levels.

## 1 Introduction

Aging, defined by the chronological passage of time since birth, is also marked by a progressive decline in physiological function and systemic resilience (de Magalhães, 2011). While chronological age is often used as a risk factor for numerous diseases, it does not fully capture the complexity of personalized aging (Wingard et al., 1988; Weinert and Timiras, 2003; de Magalhães, 2011). Although global life expectancy continues to rise, gains in health span—defined as years lived in good health—have not kept pace. This discrepancy, together with the growing burden of age-related diseases, has intensified efforts to understand the biology of aging (Idris et al., 2020; Müller et al., 2024). Emerging research now supports the view that aging is a highly personalized process, offering more profound insights into health trajectories and disease susceptibility (Kroemer et al., 2025).

Epigenetic clocks are a class of biomarkers of aging (BoA) that estimate an individual’s biological age based on their DNA methylation profiles. Among BoA, epigenetic clocks are considered among the most sensitive, reliable, and extensively studied (Salameh et al., 2020; Belsky et al., 2022; Jiang et al., 2022). Each epigenetic clock utilizes a distinct set of cytosine-phosphate-guanine (CpG) sites to predict biological age. Most epigenetic clocks are implemented as linear models, where each selected CpG site is assigned a weight based on its association with biological aging (Horvath, 2013; Hannum et al., 2013; Levine et al., 2018; Lu, A.T et al., 2019; Belsky et al., 2022). The application of epigenetic clocks in translational research to study aging and longevity is rapidly expanding. Notably, epigenetic clocks have been incorporated into randomized clinical trials to investigate the effects of lifestyle and dietary interventions on aging (García-García et al., 2024; Bell et al., 2019; Quach et al., 2017; Nielsen et al., 2022). Most existing epigenetic clocks have been developed using data from Infinium Methylation arrays, including the Illumina HumanMethylation27 BeadChip (Bibikova et al., 2009), the HumanMethylation450 BeadChip (Bibikova et al., 2011), and the MethylationEPIC BeadChip (Moran et al., 2016).

Importantly, for epigenetic clocks to serve as robust biomarkers of aging, they must generalize well across diverse populations (Horvath and Raj, 2018; Lu et al., 2019; Biomarkers of Aging Consortium et al., 2024). However, recent studies have revealed discrepancies not only between different epigenetic clocks but also in their performance across underrepresented populations, with some population-specific biases yet to be fully characterized (Chervova et al., 2024; Yusipov et al., 2024).

One potential contributor to these disparities is genomic variation. For instance, Lu et al. (2018), analyzing data from approximately 10,000 individuals, predominantly of European ancestry, identified eight genomic variants significantly associated with epigenetic age acceleration —the difference between chronological and biological age, as measured by the Hannum and Horvath pan-tissue epigenetic clocks. These findings, further supported by twin studies (Sant’Anna Barbosa Ferreira et al., 2025), suggest that genomic factors may influence DNA methylation at age-informative CpG sites. Building on this, Cruz-González et al. (2024) investigated methylation quantitative trait loci (meQTLs)—genomic variants that affect methylation levels at specific CpG sites—and assessed their prevalence within existing epigenetic clocks. Using meQTL data from diverse populations, they reported that 76% of CpG sites in the Horvath pan-tissue clock are associated with known meQTLs.

Another mechanism by which genomic variants may impact the CpG site methylation signal is through direct overlap with the cytosine or guanine base of the CpG site itself (Daca-Roszak et al., 2015; Ong and Holbrook, 2014; Chen et al., 2013). Single nucleotide polymorphisms (SNPs) are base substitutions that exhibit population-specific allele frequency distributions (Brookes, 1999). SNP-CpG site overlaps can differentiate methylation patterns at CpG sites in populations with distinct genomic backgrounds (Daca-Roszak et al., 2015). To date, no study has systematically investigated the impact of SNP-CpG site overlaps in the context of widely used epigenetic clocks. This issue is particularly relevant for genetically diverse and underrepresented populations, such as those of Arab descent, especially in epigenetic clock development efforts (Scott et al., 2016; Yusipov et al., 2024).

In this study, we aim to characterize both the qualitative and quantitative effects of SNPs on clock outputs. Specifically, we focus on common SNPs (cSNPs), whose allele frequency (AF), that is, the frequency of the rarest genomic variant, is at least 1% (0.01) in a given population. To this end, we implement three complementary analyses (**Figure 1)**, each addressing a distinct question: (1) whether cSNP-affected CpG sites are overrepresented in epigenetic clocks, and whether overlapping cSNPs are overrepresented in specific populations; (2) whether cSNP-affected CpG sites carry disproportionately greater weights in linear model-based epigenetic clocks; and (3) to what extent cSNP-induced methylation variation can alter epigenetic age estimates.

**Figure 1.**
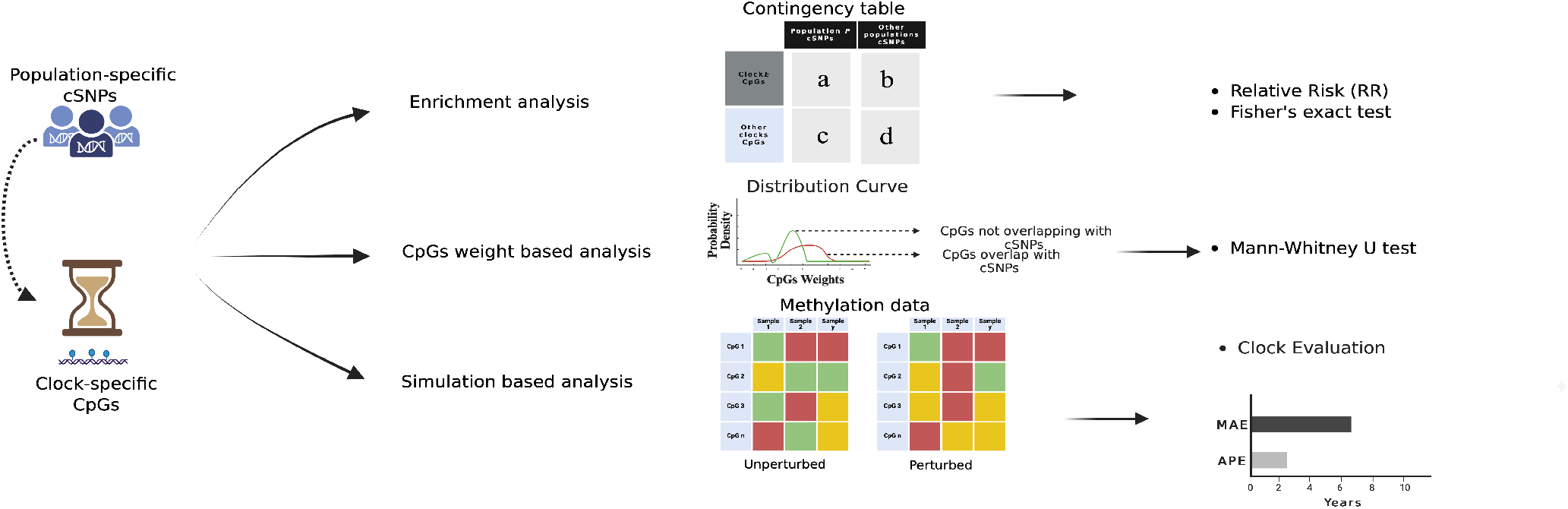
The study conceptual framework. The schema outlines the study framework, composed of three complementary analyses: (I) Enrichment analysis, where we assess cSNP-CpG overlaps enrichment across epigenetic clocks and populations through contingency table analysis; (II) CpGs weight based analysis, where we contrast the linear model weights between cSNPs-overlapping and cSNPs-non-overlapping CpG sites per clock; and (III) Simulation based analysis, where the epigenetic clocks’ performances are assessed on actual methylation data before and after altering methylation levels (i.e., perturbation) according to the presence of simulated SNP-CpG overlaps, assessed with mean absolute error (MAE) and average perturbation error (APE). See methods for details.

## 2 Method

### Selecting and preparing epigenetic clocks data

We collected the most representative epigenetic clocks based on the number of citations of their corresponding publications. Epigenetic clocks that did not disclose the list of CpG sites or sufficient information regarding their statistical model—such as GrimAge (Lu, A.T et al., 2019), DamAge (Ying, K et al., 2024), and AdaptAge (Ying, K et al., 2024)—were excluded from the study (**Table S1**).

The model type (e.g., linear vs. non-linear) was not a selection criterion; however, all selected epigenetic clocks happened to be implemented as linear models.

For each clock, we extracted the list of CpG sites identified by their array-specific probe IDs (e.g., cg25410668) and their corresponding regression weights (**Table S2**). We created clock-specific CpG site lists that contain genomic coordinate information for each clock’s CpG sites and saved them in BED files. A combined BED file of all CpG sites used across epigenetic clocks (“all clock CpGs”) was also generated. Additionally, we compiled metadata for each clock, including clock size (i.e., the number of CpG sites) and citation count (**Table S1**).

### Selecting and preparing a reference genomic dataset

We utilized the Genome Aggregation Database (gnomAD) v4.1.0 (March 2024 release). This repository includes genomic variants detected in over 730,000 whole-exome and 76,000 whole-genome profiles, aggregated at the highest level into nine genomic ancestry groups (also known as populations, **Table S3**) (Karczewski et al., 2020).

Our analysis focused exclusively on single-nucleotide polymorphisms (SNPs). For each population, we retrieved SNP genomic coordinates and variant details—including chromosome, genomic position, reference allele, alternate allele, and allele frequency (AF)—and formatted them into BED files. SNPs with AF greater than 0.01 were labelled as common SNPs (cSNPs). We then aggregated the cSNPs from all populations into a unique pan-population list (“all population cSNPs”).

All gnomAD SNP data were processed using bcftools v1.2 (Danecek et al., 202); the detailed command-line workflow is provided in **Table S4**.

### Data integration

We intersected the population-specific cSNP list with the CpG site list for each epigenetic clock, using the GRCh38 reference genome assembly. For each clock, we assessed cSNP coordinates for overlap with the genomic position of either the cytosine or guanine base of the CpG sites. The liftOver tool (Perez et al., 2025) was used to convert genomic coordinates when they were provided in the earlier GRCh37 assembly, as in the Horvath pan-tissue (Horvath, 2013) and Hannum (Hannum et al., 2013) epigenetic clocks. All genomic coordinate operations were performed using bedtools v2.26 (Quinlan and Ira, 2010). Full command-line details are in **Table S4**.

### Basic statistics

The Pearson’s test was conducted using *scipy*.*stats*.*pearsonr* Python (version 3.8) function.

### Enrichment analysis

To quantify the extent to which epigenetic clocks or populations are enriched for cSNP-CpG overlaps, we conducted two enrichment analyses:

- *Is there an epigenetic clock whose CpG sites disproportionately overlap cSNPs detected in at least one population*?

We define the background as the set of all CpG sites included in any epigenetic clock (“all clock CpGs”).

For each epigenetic clock ***E*** and population ***P***, we classify the CpG sites in “all clock CpGs” by whether they are included in ***E*** and whether they overlap with cSNPs from ***P***.

- *Is there a population whose cSNPs disproportionately overlap CpG sites used in at least one epigenetic clock*?

We defined the background as the set of all cSNPs identified in at least one population (“all population cSNPs”).

For each population ***P*** and epigenetic clock ***E***, we classify the cSNPs in “all population cSNPs” by whether they are present in ***P*** and whether they overlap with CpG sites from ***E***.

The resulting contingency tables and the calculation of enrichment scores—using relative risk (RR)—are described in **Box S1**. The statistical significance was assessed using Fisher’s exact test, and Benjamini-Hochberg false discovery rate (FDR) was used for adjusting for multiple testing (Altman, 1991; Benjamini, Y. and Hochberg, Y., 1995).

### Clock-specific CpG site weight analysis

We analyzed the weights assigned to the CpG sites of each clock to evaluate whether cSNP-CpG overlaps may influence epigenetic age estimates. For each clock ***E***, we first categorize its CpG sites into two groups, depending on whether they overlap with “all population cSNPs”. We then compared the absolute values of the linear weights between these two groups.

We used the non-parametric Mann-Whitney U test (Nachar, N., 2008) to perform the comparison, as all epigenetic clocks showed significant deviation from normality (Shapiro-Wilk test, p < 0.05) (Altman, 1991). Multiple testing correction was applied using FDR (Benjamini, Y. and Hochberg, Y., 1995).

### Simulation-based analysis

To evaluate the potential impact of cSNP-CpG overlaps on epigenetic age estimates, we performed simulation tests using publicly available DNA methylation datasets from four cohorts: Admixed American (GSE40279), African American (GSE154683), South Korean (GSE225544), and British (GSE196696). These cohorts were matched to their corresponding gnomAD ancestry groups as detailed in **Table S5**. All datasets were obtained through the Gene Expression Omnibus (GEO) (Barrett et al., 2013) and generated using either the Infinium HumanMethylation450 or Infinium MethylationEPIC array (Bibikova et al., 2011; Moran et al., 2016). We filtered all datasets to include only samples from healthy individuals.

For each methylation dataset, we used two ancestry-matched lists of SNPs to conduct three simulation tests. The first list is composed of common SNPs (AF > 0.01; cSNPs). In contrast, the second list included all identified SNPs without AF filtering (nofiltSNPs).

The simulations were conducted under two distinct frameworks that differ in how AF was applied to model the expected carrier count for a SNP in a specific population:

- *SNP-specific AF framework:* The actual AF of each SNP in the corresponding population guided the number of samples affected by that SNP during perturbation. Under this framework, we conducted **Simulation Test I** using only cSNPs.
- *Average AF framework*. The mean AF (across all overlapping SNPs within each population) was uniformly applied, simulating a more generalized perturbation scenario independent of SNP-specific AF variability. Under this framework, we conducted **Simulation Test II** using the cSNPs and **Simulation Test III** using nofiltSNPs.

Each Simulation Test was repeated ten times. In each iteration, we executed the following schema:

1. . For each SNP, we randomly selected samples, where ; *N* is the total number of healthy individuals in the dataset, and *f* is the SNP frequency. To avoid redundancy, each sample was selected only once per SNP across all ten iterations.
2. . The β-values of the CpG sites overlapping selected SNPs were perturbed to model the influence of the SNPs on methylation. We followed prior findings (Daca-Roszak et al., 2015) to determine the perturbation magnitude, which depends upon the genotype zygosity estimation (homozygous vs. heterozygous) and upon the Hardy-Weinberg Equilibrium assumptions (Hartl and Clark, 2007).

The complete simulation algorithm is provided in **Box S1**. Baseline biological ages were first predicted from the unperturbed DNA methylation datasets using all considered epigenetic clocks. Subsequently, at each iteration, biological ages were re-estimated from the perturbed datasets using the same models. All age predictions were computed with the Biolearn Python library (Ying, K et al., 2024).

For each dataset and clock, we then computed the following metrics:

- Mean Absolute Error (MAE): is the difference between chronological age and biological age predicted in the original sample (referred to as the “baseline error”).
- Average Perturbation Error (APE): is the difference between biological age predicted from the unperturbed and perturbed datasets, averaged across all 10 iterations for each sample.

The scripts used for simulations and CpGs weight analyses are provided in the Study GitHub Repository.

#### 3 Results

### Selected epigenetic clocks and populations

We analyzed representative sets of epigenetic clocks, human populations, and their corresponding common single nucleotide polymorphisms (cSNPs) to evaluate the impact of cSNPs on epigenetic clock estimations.

The selected epigenetic clocks included Horvath pan-mammalian (Lu et al., 2023), PhenoAge (Levine et al., 2018), DunedinPACE (Belsky et al., 2022), Horvath pan-tissue (Horvath, 2013), Horvath skin (Horvath et al., 2018), Hannum (Hannum et al., 2013), Xiaohui Wu (Wu et al., 2019), Chunxiao Li (Li et al., 2018), and PedBE (McEwen et al., 2020) (**Table S1**). All chosen epigenetic clocks were implemented as linear models, which estimate age by applying weighted coefficients to methylation levels (from 0 to 1) at specific CpG sites (age predictors).

We considered nine major genomic ancestry groups (populations) represented in gnomAD v4.1.0: African (AFR), Amish (AMI), Admixed American (AMR), Ashkenazi Jewish (ASJ), East Asian (EAS), Finnish (FIN), Middle Eastern (MID), non-Finnish European (NFE), and South Asian (SAS). Population-specific cSNP counts are provided in **Table S3**; AFR carried the largest number of cSNPs, followed by MID and AMR.

### cSNPs–CpG site overlap and enrichment patterns in epigenetic clocks and populations

We first identified overlaps between cSNPs and clock-specific CpG sites. A cSNP-CpG overlap was defined as a cSNP intersecting either the cytosine or guanine base of a clock-specific CpG site. Across all 81 population-clock combinations, 14 showed no overlaps, and the average number of overlaps per combination was small (mean = 3; see Methods and **Figure S1**). When comparing population size with the number of cSNP-CpG overlaps, no significant association was observed (r = 0.33, p = 0.4654). By contrast, the size of the epigenetic clock (i.e., the number of CpG sites included) was strongly associated with the number of cSNP-CpG overlaps (r = 0.95, p = 0.0001; **Figure S2**).

To evaulate whether a specific epigenetic clock or population is disproportionately affected by the overlaps, we conducted two complementary enrichment analyses at the most granular level: (i) per epigenetic clock, testing whether CpG sites overlapping cSNPs were enriched in specific populations; and (ii) per population, testing whether cSNPs were preferentially enriched in specific epigenetic clocks.

In the first analysis, for each epigenetic clock, we used as background the set of all CpG sites included in any epigenetic clock (“all clock CpGs”; see Methods). None of these enrichments were statistically significant after multiple-testing correction (FDR < 0.05). Interestingly, the Chunxiao Li clock exhibited the highest average enrichment across populations (relative risk (RR) = 2.6), followed by Hannum (RR = 1.4), and PhenoAge (RR = 1.2) (**Figure 2(A)**, upper panel).

**Figure 2.**
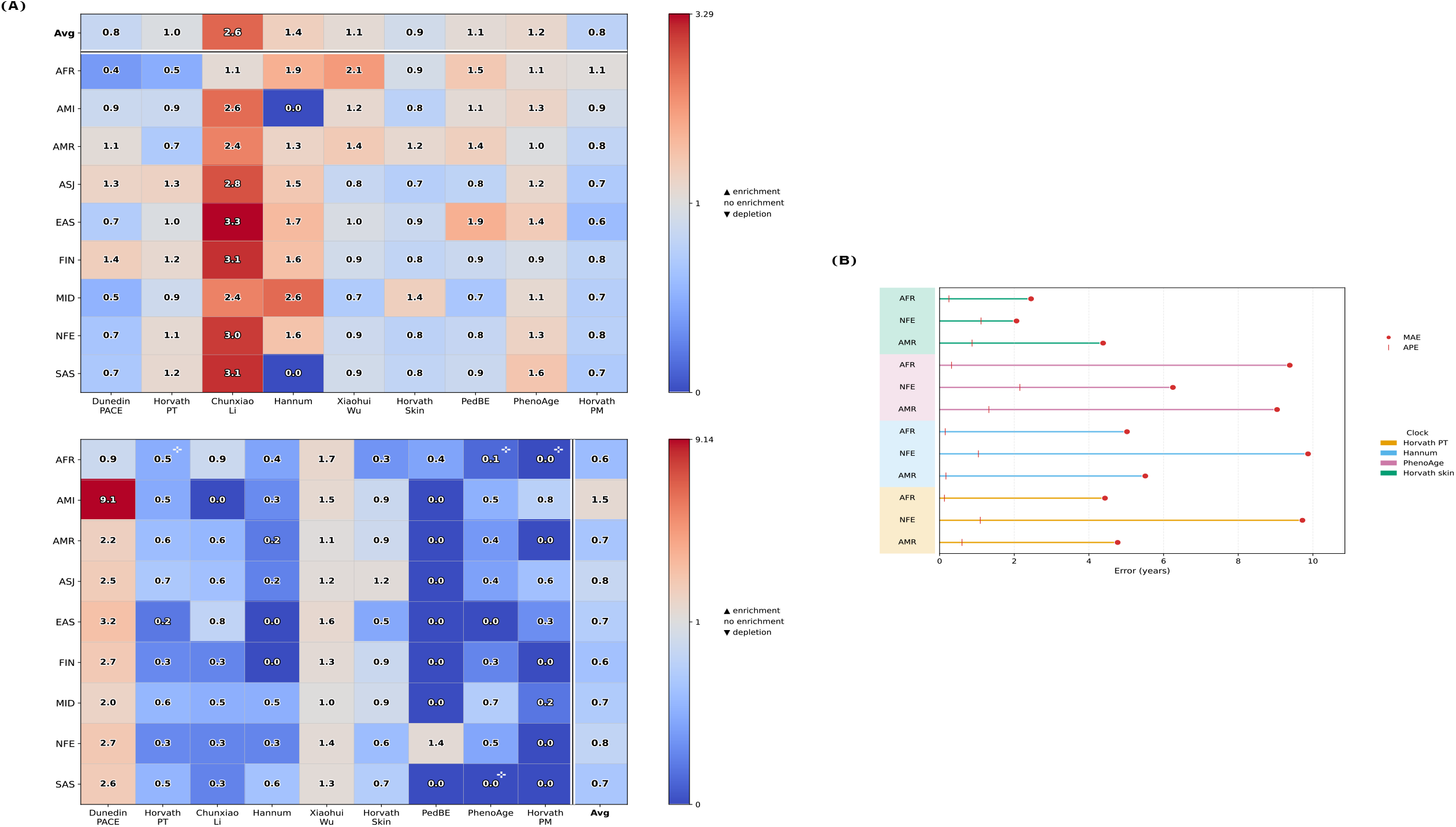
Summary of analyses assessing cSNP-CpG overlaps in epigenetic clocks. **Panel A:** Enrichment analysis of cSNP-CpG overlaps across epigenetic clocks and populations. The upper heatmap displays enrichment scores (relative risk, RR) for each clock, with the “**Avg**” column representing the mean enrichment across populations. The lower heatmap shows enrichment scores by population, with the “**Avg**” row indicating the mean enrichment across epigenetic clocks. Color scales are centered at RR = 1, denoting depletion, no enrichment, or enrichment. ^*^ Horvath PT: Horvath pan-tissue; Horvath PM: Horvath pan-mammalian. **Panel B:** Simulation-based performance evaluation of epigenetic clocks. Bar plots show mean absolute error (MAE;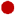) and average perturbation error (APE; |) for each clock-population combination, based on Simulation Test III (all SNPs). Results are presented for three DNA methylation datasets representing distinct populations: Admixed American (AMR), Non-Finnish European (NFE), and African (AFR). Horvath PT: Horvath pan-tissue.

In the second analysis, for each population, we used as background all cSNPs identified in at least one population (all population cSNPs). Four significant population-clock associations (FDR < 0.05) were observed: AFR showed significant enrichment in the PhenoAge, Horvath pan-mammalian, and Horvath pan-tissue epigenetic clocks, whereas SAS showed significant enrichment in the PhenoAge clock. In this analysis, enrichment levels varied across epigenetic clocks but remained generally consistent across populations, with only a few notable exceptions (e.g., AFR–DunedinPACE; **Figure 2(A)**, lower panel).

Overall, enrichment scores varied across epigenetic clocks and populations, with only limited cases reaching statistical significance. The small number of cSNP-CpG overlaps likely reduced statistical power, constraining our ability to detect robust effects. Nonetheless, the observed enrichment patterns suggest potential susceptibility of certain epigenetic clocks to cSNP-related biases and underscore the need for deeper quantification of their impact on biological age estimation.

### Impact of cSNP–CpG site overlaps on age estimates: clock-specific CpG weight analysis

The first step in quantifying the effects of cSNP-CpG overlaps was to investigate the weights assigned to CpG sites in each epigenetic clock’s linear model. Because overlap counts at the single-population level were too small or absent, cSNPs were pooled across all populations. For each clock, we then compared the distributions of absolute linear regression weights between overlapping and non-overlapping CpG sites. Across all epigenetic clocks, no statistically significant differences were observed (FDR < 0.05; **Table 1**).

**Table 1:**
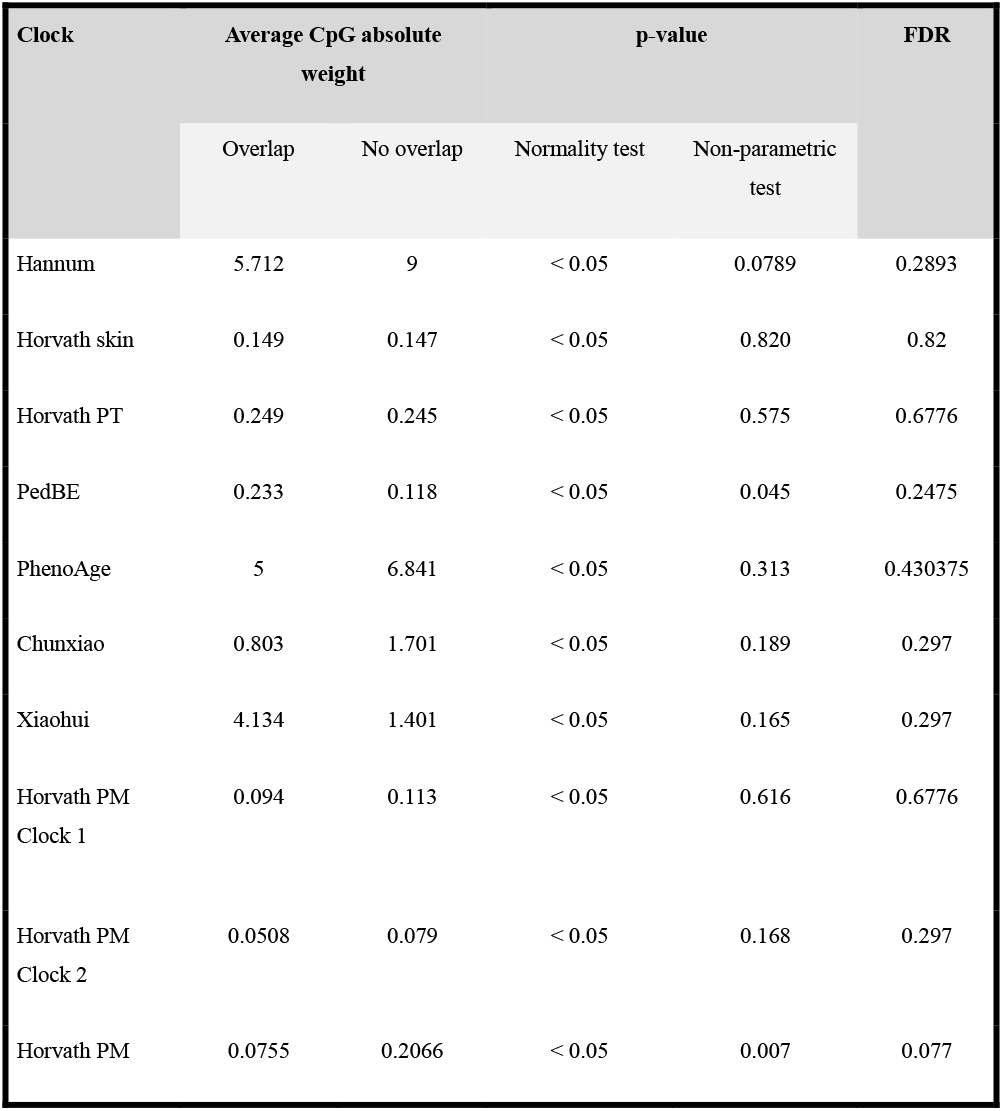
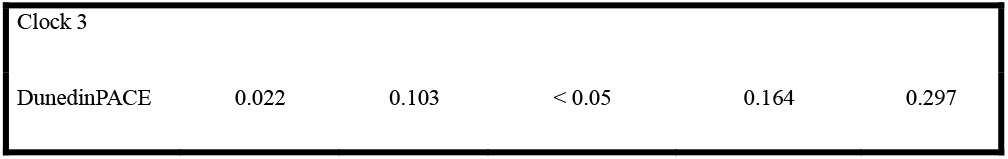
Epigenetic clock CpG weight analysis results. The table reports the mean of weights of two CpG site categories: CpGs that overlap with cSNPs and those without overlaps. Normality was assessed using the Shapiro-Wilk test. Differences between distributions were evaluated using the Mann-Whitney U test, and multiple testing correction was applied using the Benjamini-Hochberg false discovery rate (FDR). Horvath PT: Horvath pan-tissue; Horvath PM: Horvath pan-mammalian.

| Clock | Average CpG absolute weight |  | p-value |  | FDR |
| --- | --- | --- | --- | --- | --- |
|  | Overlap | No overlap | Normality test | Non-parametric test |  |
| Hannum | 5.712 | 9 | < 0.05 | 0.0789 | 0.2893 |
| Horvath skin | 0.149 | 0.147 | < 0.05 | 0.820 | 0.82 |
| Horvath PT | 0.249 | 0.245 | < 0.05 | 0.575 | 0.6776 |
| PedBE | 0.233 | 0.118 | < 0.05 | 0.045 | 0.2475 |
| PhenoAge | 5 | 6.841 | < 0.05 | 0.313 | 0.430375 |
| Chunxiao | 0.803 | 1.701 | < 0.05 | 0.189 | 0.297 |
| Xiaohui | 4.134 | 1.401 | < 0.05 | 0.165 | 0.297 |
| Horvath PM<br>Clock 1 | 0.094 | 0.113 | < 0.05 | 0.616 | 0.6776 |
| Horvath PM<br>Clock 2 | 0.0508 | 0.079 | < 0.05 | 0.168 | 0.297 |
| Horvath PM | 0.0755 | 0.2066 | < 0.05 | 0.007 | 0.077 |
| Clock 3 |  |  |  |  |  |
| DunedinPACE | 0.022 | 0.103 | < 0.05 | 0.164 | 0.297 |

These findings suggest that cSNP-CpG overlaps are not preferentially associated with the most influential features of the clock models. However, this does not exclude the possibility that overlapping CpGs could still contribute additional effects on biological age estimation.

### Impact of SNP–CpG site overlaps on age estimates: simulation analysis

Finally, we aimed to computationally quantify the impact of SNP-CpG site overlaps on epigenetic clock predictions through simulation-based analyses. For this purpose, we used four publicly available DNA methylation datasets, each aligned with a specific gnomAD population (AMR, NFE, AFR, and EAS; see **Table S5** and Methods). We excluded epigenetic clocks not originally designed to predict biological age in adults, with limited methylation data availability, or lack of a standardized implementation framework: Horvath pan-mammalian, Chunxiao Li, Xiaohui Wu, DunedinPACE, and PedBE. Specifically, the Xiaohui Wu and PedBE epigenetic clocks are pediatric-specific; the pan-mammalian clock is trained on a specialized mammalian methylation array with limited applicability to human samples (Arneson et al., 2022); DunedinPACE estimates the pace of aging rather than chronological age; and the Chunxiao Li clock is not supported by standard R/Python packages.

For each methylation dataset, two SNP lists were considered: common SNPs filtered by allele frequency (AF > 0.01; denoted cSNPs) and SNPs without AF filtering (denoted nofiltSNPs) (see Methods). For each dataset, β-values at SNP-overlapping CpG sites were perturbed *in silico* to mimic SNP-driven effects on methylation, following previously established patterns (Daca-Roszak et al., 2015).

We performed three simulation tests, each run for 10 iterations per dataset, varying the number of affected CpGs and SNP inclusion criteria. **Simulation Test I** modeled the effects of cSNPs by estimating, for each individual, the probability of including the effect of a cSNP based on its population-specific AF. **Simulation Test II** applied a stronger perturbation by using global mean AF values rather than cSNP-specific AF, thereby incorporating the effects of less frequent cSNPs. **Simulation Test III** further stressed the system by following the Test II approach but expanding the candidate set to include non-common SNPs (nofiltSNPs). In summary, these three simulation tests represent progressively increasing levels of perturbation strength, with Test I reflecting common variation and Tests II–III exploring the impact of rarer and broader SNP effects.

For each case, the perturbation effect was quantified as the mean difference between predictions from unperturbed and perturbed data (**Box S1**). To complete the analysis, we computed first the average perturbation error (APE) as the difference between predicted biological ages before and after perturbation,. For the perturbed estimation, the mean across the 10 iterations was computed for each sample, yielding a single average predicted age per individual. As expected, the largest deviations were observed in **Simulation Test III**, with APE values ranging from 0.1 to 2.1 years (**Table S6**). By comparison, narrower APE ranges were found in **Simulation Test I** (0-1.1 years) and **Simulation Test II** (0-0.2 years).

Because the impact of the APE depends on the original distance between biological age and chronological age, we compared APE with the mean absolute error (MAE), defined as the difference between chronological age and biological age predicted in the original data. In all cases, APE values were consistently smaller than MAE, indicating that although SNP-CpG overlaps introduced measurable deviations, their magnitude was modest relative to the epigenetic clocks’ baseline error (**Figure 2(B)**).

Overall, the simulations demonstrate that SNP-CpG overlaps can affect epigenetic age predictions, particularly in scenarios with a high level of SNP-induced variation. However, the extent of deviation remains smaller than typical model error, possibly suggesting a limited impact on real-world applications across diverse populations.

## 4 Discussion

Epigenetic clocks are biomarkers of aging based on DNA methylation patterns, useful for studying health and disease (Salameh et al., 2020; Belsky et al., 2022; Jiang et al., 2022). Yet, genomic variants can distort methylation signals: cytosine-overlapping SNPs abolish methylatable sites, while guanine-overlapping SNPs disrupt probe chemistry, adding noise (Daca-Roszak et al., 2015). In this study, we assessed whether common SNPs (cSNPs) overlapping CpG sites affect the performance of widely used epigenetic clocks across nine populations, including Middle Eastern (MID) and Amish (AMI) ancestries, which have been largely underrepresented in previous reseach.

We found little evidence of significant cSNP-CpG overlaps, likely reflecting quality control that excludes such sites during clock development (Burrage et al., 2019; Lu et al., 2023). Weight-based analyses of regression models showed no significant differences between overlapping and non-overlapping CpG sites, suggesting minimal influence on predictions. Simulations-based perturbation tests further confirmed that even biased overlap scenarios introduced discrepancies smaller than typical clock error rates.

Limitations in our analysis include skewed sample sizes across gnomAD populations (e.g., 37,000 African vs. ∼3,000 Middle Eastern genomes), heterogeneous “healthy” definitions across publicly available metahylation datasets, and a lack of matched genomic-methylation data from the same individuals. Nonetheless, our results align with studies showing that while genomic variants, such as Telomerase Reverse Transcriptase (TERT), can influence age acceleration (Lu et al., 2018; Lin, 2021), direct cSNP-CpG overlaps add negligible noise (Ong & Holbrook, 2014; Chen et al., 2013).

Despite these limitations, the errors from cSNP-CpG overlaps were consistently small, supporting robust clock performance across diverse ancestries. However, we observed wide MAE variation (2.0–9.8 years) across healthy cohorts (**Figure 2(B)**). Future work should address other drivers of variability—environmental, socioeconomic, or genetic—to improve clinical and global utility of epigenetic aging models.

## Data Availability

All data used in this study are publicly available. Genomic variant data were obtained from the gnomAD Browser. DNA methylation datasets (GSE40279, GSE154683, GSE225544, GSE196696) are accessible through the Gene Expression Omnibus (GEO).

https://gnomad.broadinstitute.org/

https://www.ncbi.nlm.nih.gov/geo/

## 6 Author Contributions

DG: Conceptualization, writing–review and editing, formal analysis, supervision. VL: Writing– review and editing, formal analysis, supervision. JT: Writing–review and editing, supervision. EH: Writing–review and formal analysis. TS: Data curation, investigation, formal analysis, and writing– original draft. All authors contributed to reading and approving the final manuscript.

## 8. Acknowledgments

The authors thank King Abdullah University for Science & Technology (KAUST) Supercomputing Laboratory for providing computational resources essential for the analysis.

## 10 Supplementary Material

Supplementary tables and figures can be found in Supplementary_file_1.docx and Supplementary_Tables.xlsx.

## 12 Data Availability Statement

All data used in this study are publicly available. Genomic variant data were obtained from the gnomAD Browser. DNA methylation datasets (GSE40279, GSE154683, GSE225544, GSE196696) are accessible through the Gene Expression Omnibus (GEO). Custom code developed for simulations and CpG weight analyses is openly available in the Study GitHub Repository.

