## Supplementary File1 for "Epigenetic Age Prediction Remains Stable Across Common Variants and Diverse Ancestries"

**Supplementary File 1**

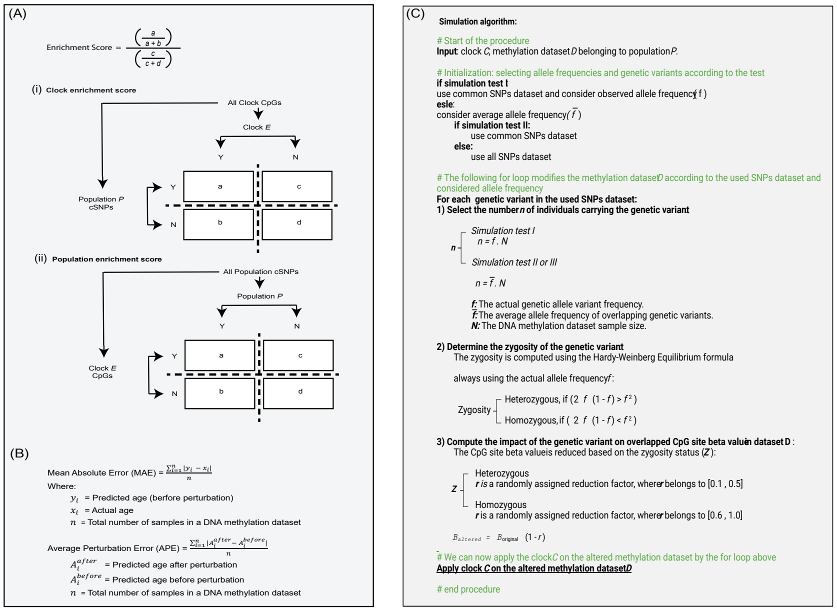


Box S1: Mathematical formulas and simulation algorithm used in this study.

(A) Enrichment score formulas and contingency tables. Schematics illustrate the calculation of (i) clock enrichment scores, defined as the relative risk (RR) of cSNP-CpG site overlaps occurring within a given epigenetic clock compared to all other clocks, and (ii) population enrichment scores, defined as the relative risk (RR) of overlaps within a specific population compared to all other populations (see Methods for details).

(B) Evaluation metrics.

- *Mean Absolute Error (MAE):* the average absolute difference between predicted age (before perturbation) and chronological age across all samples.
- *Average Perturbation Error (APE):* the average difference between unperturbed and perturbed age predictions, computed as the mean of 10 simulation iterations per sample.

(C) Simulation algorithm. Flowchart depicting the perturbation-based simulation procedure applied to SNP-overlapping CpG sites.

Figures:

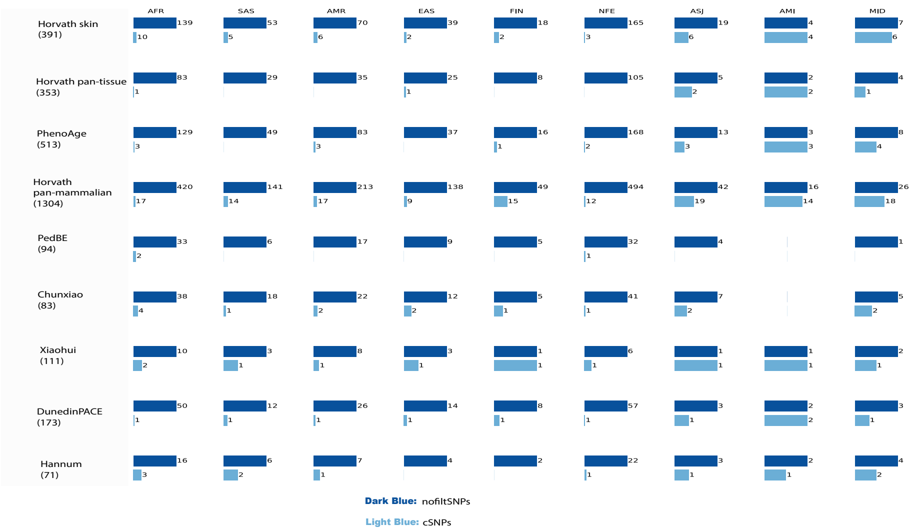


Figure S1: SNP-CpG overlap counts across clocks and populations.

Bar plot showing the SNP-CpG site overlap counts for each clock–population pair. "nofiltSNPs" include every SNP identified in a given population, regardless of allele frequency (AF). "cSNPs" are defined as those with AF > 1% (0.01) in that population. Bar heights are scaled independently within each clock-population pair; therefore, comparisons should be made within pairs rather than across different pairs.

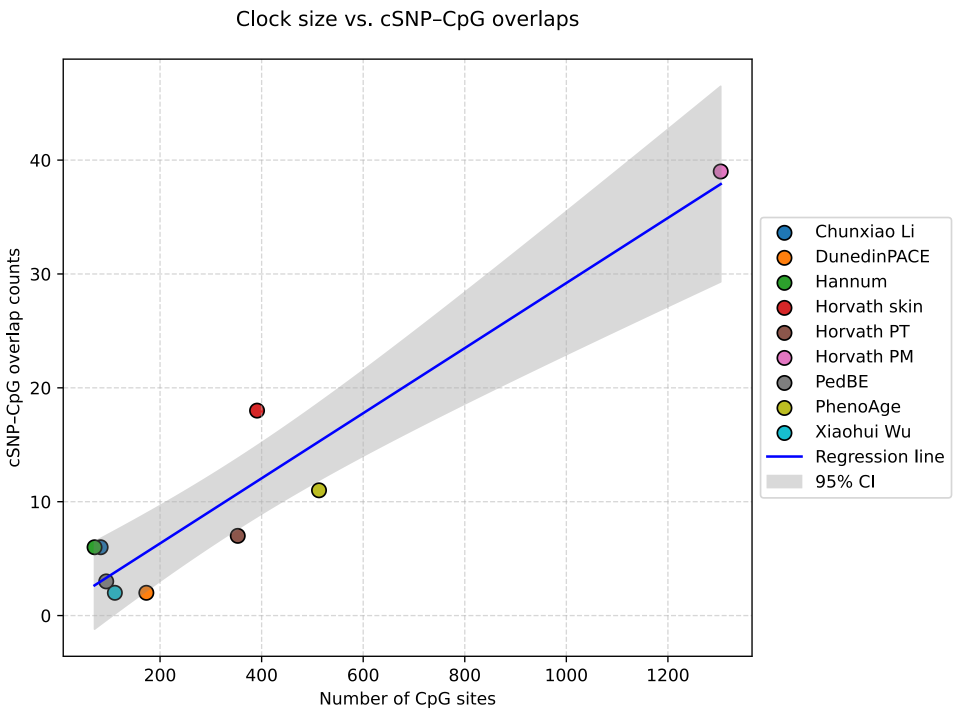


Commented [VL1]: I cropped the image

Figure S2: Correlation analysis between the epigenetic clock size and cSNP-CpG counts.

Scatter plot showing the relationship between epigenetic clock size (i.e., the number of CpG sites used as age predictors) and the cSNP-CpG site overlap counts. Each point represents a clock, with labels identifying specific clocks. A strong positive Pearson's correlation ($r = 0.95$, $p = 0.0001$). The blue regression line represents the linear trend, with a 95% confidence interval (shaded area). The statistical test was done using *scipy.stats.pearsonr* in Python. Horvath PT: Horvath pan-tissue; Horvath PM: Horvath pan-mammalian.
